# AI-Driven Precision Nutrition for Predicting Iron Deficiency Using Genomics, Polygenic Risk Scores, Dietary Patterns, and Personalized Dietary Recommendations

**DOI:** 10.64898/2026.07.06.26357351

**Authors:** B Shravani, S Vijetha, Yashna Bhandary, Vijayalaxmi, Sanjana R Otihal

**Author notes:** Principal corresponding author.

## Abstract

**Background:** Iron Deficiency Anemia (IDA) is one of the most prevalent nutritional disorders globally and a leading cause of Disability Adjusted Life Years (DALYs). Conventional diagnostic methods fail to detect deficiencies at an early stage and rarely account for individual genetic predisposition.

**Methods:** This study proposes an end-to-end AI-driven precision nutrition pipeline integrating public Genome-Wide Association Study (GWAS) data and NHANES phenotypic data encompassing demographics, dietary intake, anthropometrics, and hematology. A synthetic genotype matrix was simulated for 400 GWAS-filtered SNPs using Hardy-Weinberg Equilibrium. Data preprocessing included missing value imputation, feature engineering, and SMOTE class balancing. Four machine learning models namely, Logistic Regression, Random Forest, Artificial Neural Network (ANN), and XGBoost were implemented and evaluated for both IDA classification and haemoglobin regression tasks.

**Results:** XGBoost achieved state-of-the-art performance with ROC-AUC = 0.9981 for classification and *R*^2^ = 0.9903 for haemoglobin prediction. Polygenic Risk Score (PRS) stratification classified participants into low (73%), moderate (18%), and high (9%) risk tiers. Pathway burden analysis identified the Hepcidin Regulation pathway as the highest burden pathway in high-risk individuals.

**Conclusion:** The integration of genomics, machine learning, and nutritional science through a Pathway-Burden Precision Nutrition Engine produced gene-specific, evidence-graded dietary recommendations, demonstrating significant potential for early and personalised IDA prevention.

## Introduction

Iron deficiency anemia (IDA) is the most common form of malnutrition disorder globally and one of the main causes of anemia-related disability-adjusted life years (DALYs). In the Global Burden of Disease 2021 survey, 34.5 million DALYs were reported due to iron deficiency, with most of the burden falling on South Asia [1]. Specifically in India, the National Family Health Survey (NFHS-5, 2019-2021) reported that 57.0% of women of reproductive age, 52.2% of pregnant women, and 67.1% of children aged 6-59 months are affected by anaemia [2]. IDA is responsible for 15-20% of maternal mortality directly and 45-50% indirectly, and childhood IDA results in 1.3% of India’s GDP loss [2].

Despite large-scale interventions such as the Anaemia Mukt Bharat programme serving approximately 650 million people since 2018, anaemia prevalence among adolescents has increased from 54.0% to 59.2% between NFHS-4 and NFHS-5. A meta-analysis involving 157 studies from India showed prevalence ranging from 15.9% to 83.9% across states [3].

Diet plays an enormous role in IDA aetiology. More than 95% of iron in the Indian diet originates from non-heme plant sources, with bioavailability of only 2-10% compared to 25-30% for heme iron [4]. High phytate intake (1,287-2,500 mg/day) may reduce iron bioavailability up to five-fold, while tea polyphenols further inhibit absorption. Vitamin C enhances non-heme iron bioavailability, though consumption levels vary considerably across the population.

The genetic architecture of IDA adds another layer of complexity. A GWAS meta-analysis of 257,953 subjects identified 123 loci influencing iron-related phenotypes, including key variants in *TMPRSS6, HFE, TF*, and *SLC40A1* [5]. Variant rs855791 of *TMPRSS6* significantly increases IDA risk, while rs1800562 (C282Y) of *HFE* is rarer in Asian subjects, underscoring the need for population-specific analysis. Despite the emergence of Polygenic Risk Scores (PRS) for personalised risk estimation, approximately 78% of GWAS samples used in existing PGS models are of European descent. A 2024 multi-ancestry study demonstrated approximately 51.3% decreased prediction accuracy when applying European-derived PRS to non-European populations [6].

Machine learning (ML) has shown considerable promise in IDA prediction. XGBoost models have attained AUC scores up to 0.814 in iron deficiency screening and 96.95% accuracy in anaemia classification among Indian populations [7]. However, no prior study has combined bioavailability-corrected dietary iron features with GWAS-derived PRS and representative national phenotypic datasets in a unified predictive framework. This study addresses that gap.

### Literature Review

#### Machine Learning for Iron Deficiency and Anaemia Prediction

Pullakhandam and McRoy [7] demonstrated that ML algorithms based on complete blood count (CBC) parameters can diagnose IDA accurately and outperform rule-based methods, though without considering genetic or nutritional factors. Qasrawi et al. [8] applied ML to study associations between dietary intake and anaemia in female students, confirming dietary habits as significant predictive features, though genetic predisposition was not explored. Darshan et al. [9] developed explainable AI (XAI) models to differentiate IDA from aplastic anaemia using blood parameters, improving clinical transparency but remaining limited to diagnosis without prevention focus.Tepakhan et al. [10] applied Random Forest and Gradient Boosting to distinguish IDA from thalassemia, achieving strong results but confined to CBC features.

#### Genomics and Polygenic Risk Scores

Kim et al. [11] presented a Stacked Neural Network Polygenic Risk Score (SNPRS) method superior to conventional PRS approaches for complex disease prediction. Zhao et al. [12] introduced the PUMAS-ensemble framework, demonstrating that ensemble-based PRS consistently outperforms individual scores in disease risk stratification. Zhuang et al. [13] showed that incorporating functional genomic annotations during PRS construction helps prioritize biologically relevant variants. Moreno-Grau et al. [14] investigated PRS portability across ancestries, identifying significant performance differences that motivate population-specific recalibration.

#### Integration of Genomic and Clinical Data Using AI

Hrytsenko et al. [15] found that combining PRS with clinical features substantially improves blood pressure prediction over either variable alone. Shin [16] demonstrated a combined dietary, clinical, and genome-wide PRS model for predicting metabolic syndrome, directly informing the multimodal design of the present framework.

## Materials and Methods

### Overall Pipeline Architecture

The proposed AI-driven precision nutrition pipeline comprises an 11-step sequence operating in two modes: (i) Cohort Mode, processing NHANES phenotypic and GWAS genomic data to train population-level ML models; and (ii) Single-Patient Mode, accepting an individual’s VCF genotype file, phenotypic data, and dietary records to generate a personalised anaemia risk prediction and precision nutrition report.

Fig 1 shows the overall system architecture, and Table 1 summarises each pipeline step in detail.

**Fig 1.**
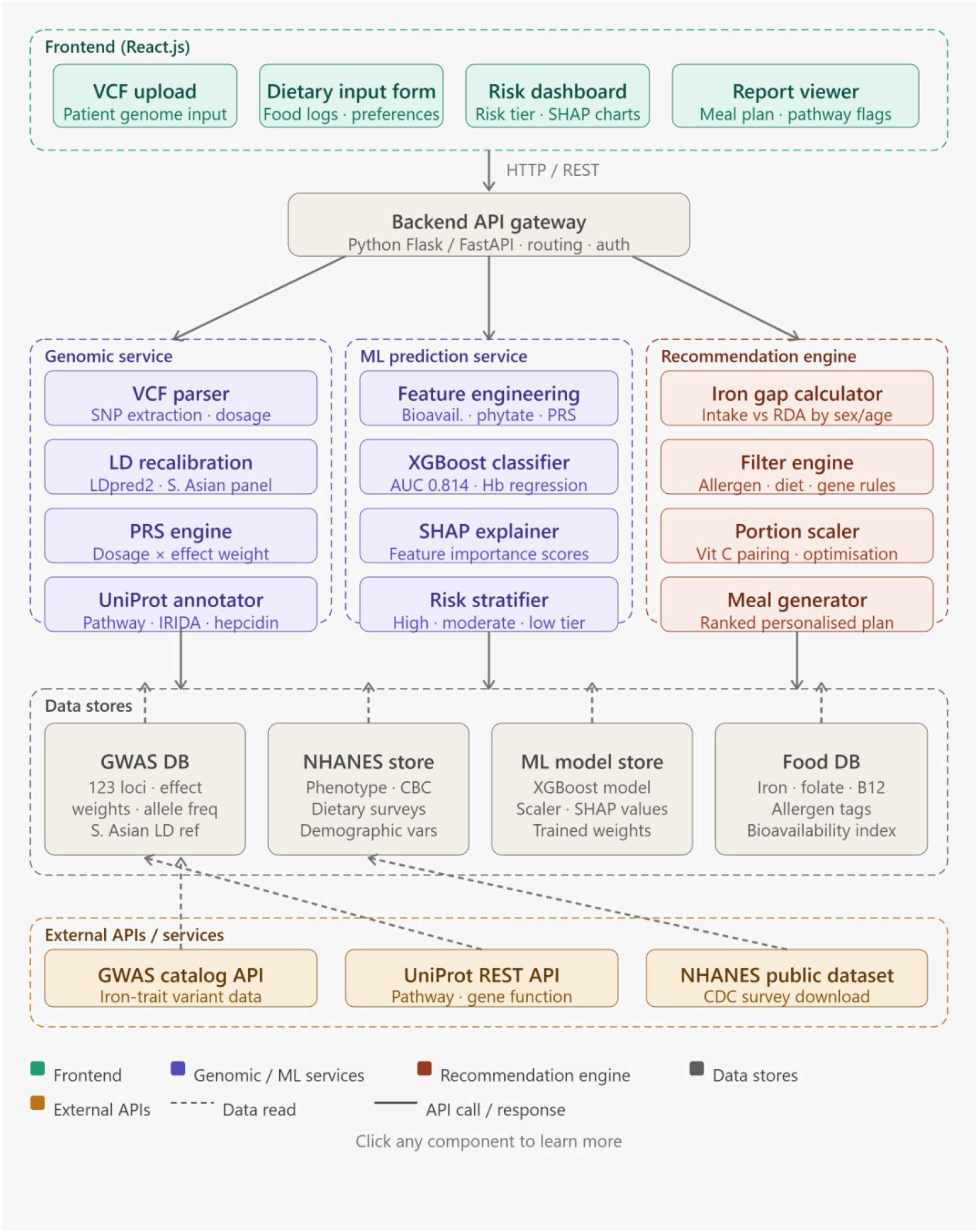
Overall architecture of the proposed AI-based personalized nutrition recommendation system for iron deficiency support. The framework integrates genomic analysis, machine learning-based risk prediction, explainable AI (SHAP), and a recommendation engine with nutritional and genomic databases to generate personalized dietary interventions based on an individual’s genomic and phenotypic profile.

**Table 1.**
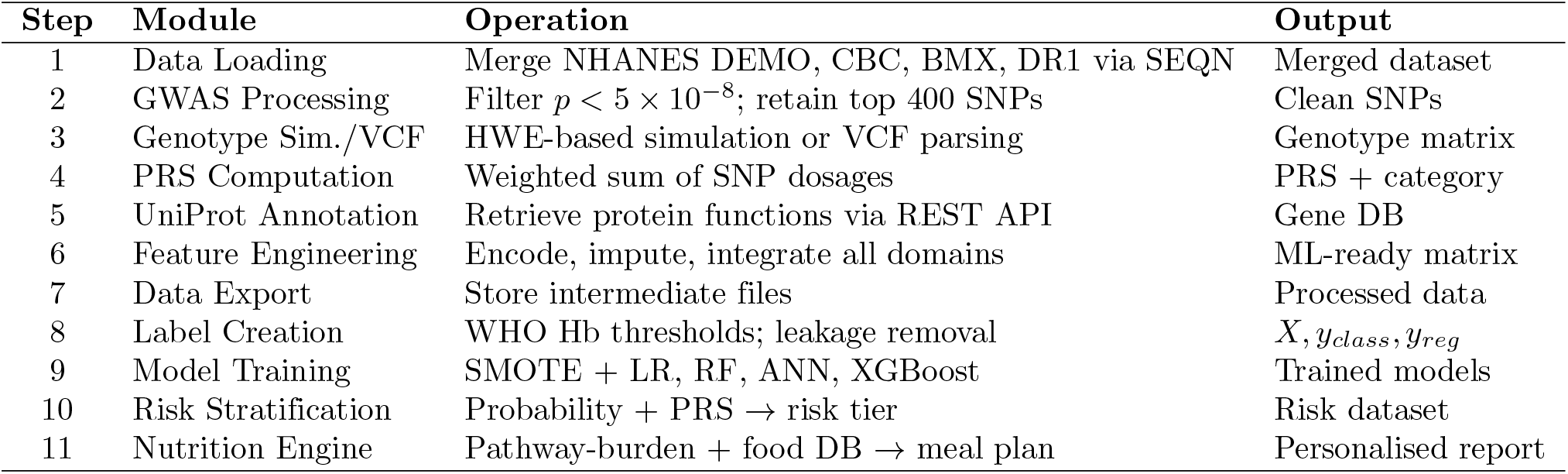
Pipeline Architecture — Step-by-Step Design.

| Step | Module | Operation | Output |
| --- | --- | --- | --- |
| 1 | Data Loading | Merge NHANES DEMO, CBC, BMX, DR1 via SEQN | Merged dataset |
| 2 | GWAS Processing | Filter $p < 5 \times 10^{-8}$; retain top 400 SNPs | Clean SNPs |
| 3 | Genotype Sim./VCF | HWE-based simulation or VCF parsing | Genotype matrix |
| 4 | PRS Computation | Weighted sum of SNP dosages | PRS + category |
| 5 | UniProt Annotation | Retrieve protein functions via REST API | Gene DB |
| 6 | Feature Engineering | Encode, impute, integrate all domains | ML-ready matrix |
| 7 | Data Export | Store intermediate files | Processed data |
| 8 | Label Creation | WHO Hb thresholds; leakage removal | $X, y_{class}, y_{reg}$ |
| 9 | Model Training | SMOTE + LR, RF, ANN, XGBoost | Trained models |
| 10 | Risk Stratification | Probability + PRS → risk tier | Risk dataset |
| 11 | Nutrition Engine | Pathway-burden + food DB → meal plan | Personalised report |

### Data Sources

#### GWAS Catalog

A search was conducted for seven iron-related traits: ferritin, haemoglobin concentration, IDA, mean corpuscular haemoglobin (MCH), serum iron, total iron binding capacity (TIBC), and transferrin saturation [5]. Quality control filters applied were: *p <* 5 ×10^*™*8^, elimination of duplicates, *β >* 0.0001, and 0.01 ≤ RAF≤ 0.99, followed by selection of the top 400 SNPs.

#### NHANE

Four CSV domains were merged on the unique participant identifier SEQN: DEMO, CBC/Lab, BMX, and DR1 [17]. Key variables included haemoglobin (LBXHGB), MCV (LBXMCVSI), MCH (LBXMCHSI), and dietary iron intake (DR1TIRON, mg/day), among others.

### Genotype Simulation via Hardy-Weinberg Equilibrium

Since NHANES does not include individual genotype data, a synthetic genotype matrix was constructed following Hardy-Weinberg Equilibrium (HWE). For each SNP with risk allele frequency *p*, genotype frequencies are:

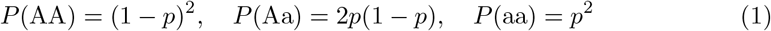

Genotype dosages (0, 1, or 2) for each of 400 SNPs were sampled for each participant according to Eq (1).

### Polygenic Risk Score Computation

The Polygenic Risk Score (PRS) for individual *i* quantifies cumulative genetic predisposition:

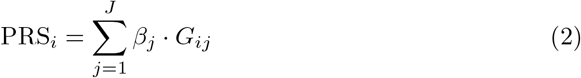

where *β*_*j*_ is the GWAS effect size for SNP *j, G*_*ij*_ ∈ {0, 1, 2} is the genotype dosage, and *J* = 400. PRS was computed as the matrix product of the genotype matrix and effect size vector. Participants were stratified as: low risk (*<*25th percentile), moderate risk (25–75th percentile), and high risk (*>*75th percentile) [19].

### Feature Engineering and Label Creation

The feature matrix comprised: demographic factors (age, sex, ethnicity, income), anthropometric measures (BMI, waist circumference), haematological indices (MCV, MCH, RDW, red blood cell count), nutritional features (iron, vitamin C, protein, calcium, total energy), PRS and its encoded category, and 400-SNP genotypes. Missing numerical values were imputed using median imputation; categorical variables underwent label encoding.

Binary anaemia labels were assigned using WHO sex-specific thresholds: IDA positive (*y* = 1) if haemoglobin *<* 12.0 g/dL (females) or *<* 13.0 g/dL (males). Continuous haemoglobin (LBXHGB) was retained as a separate regression target *y*_reg_. SMOTE [18] was applied exclusively to the training subset to prevent data leakage.

### Machine Learning Models

Four modelling configurations were evaluated on an 80/20 stratified split with SMOTE augmentation applied to training data only:

- **Logistic/Linear Regression:** L2-regularised baseline (`class weight=‘balanced’`, `max iter=1000`); StandardScaler preprocessing.
- **Random Forest:** 300 trees, `max depth=10`; feature importance via Gini impurity gain.
- **ANN (MLP):** Architecture 128–64 hidden neurons, ReLU activation, Adam optimiser, `max iter=200`; permutation importance (*n* repeats = 5).
- **XGBoost:** 300 estimators, early stopping at round 30, `learning rate=0.05`, `max depth=6`, `subsample=0.8`, `colsample bytree=0.8`, `reg alpha=0.5`.

Classification performance was measured using accuracy, precision, recall, F1-score, and ROC-AUC. Regression performance was evaluated using MAE, RMSE, and *R*^2^.

### Overall Risk Stratification Logic

A rule-based stratification system combined ML-predicted anaemia probability with PRS category:

- **High Risk:** Probability *>* 0.70, or probability *>* 0.50 with High PRS.
- **Moderate Risk:** Probability *>* 0.40, or High PRS with lower model probability.
- **Low Risk:** All remaining individuals.

### Pathway-Burden Precision Nutrition Engine

Risk-associated SNPs were mapped to iron-related biological pathways using the UniProt Knowledgebase REST API [20]. Pathway burden for pathway *k* was computed as:

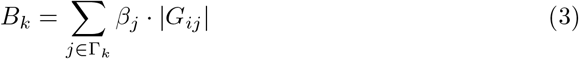

where Γ_*k*_ denotes the set of SNPs associated with pathway *k*. Pathways with the highest burden scores were mapped to a curated nutrigenomics database to generate gene-specific, evidence-graded dietary recommendations extending beyond generic iron supplementation advice.

### Theoretical Background

#### Iron Metabolism and Its Genetic Architecture

Iron is a vital micronutrient required for haemoglobin synthesis, oxygen transport, DNA synthesis, mitochondrial electron transport, and enzymatic reactions. Dietary iron absorption occurs primarily in the duodenum. Heme iron from meat is absorbed directly by enterocytes, while non-heme iron must first be reduced from Fe^3+^ to Fe^2+^ by DcytB (encoded by *CYBRD1*) and then transported via DMT1 (*SLC11A2*).

Systemic iron homeostasis is governed by the hepcidin–ferroportin axis. Hepcidin (*HAMP*) binds ferroportin (*SLC40A1*), triggering its degradation and thereby inhibiting iron efflux from enterocytes and macrophages into circulation. *HJV* positively regulates hepcidin transcription, while *TMPRSS6* (Matriptase-2) suppresses hepcidin synthesis. Loss-of-function defects in *TMPRSS6* lead to elevated hepcidin and Iron Refractory Iron Deficiency Anemia (IRIDA). Iron is transported in blood bound to transferrin (TF), taken up via transferrin receptors (TFRC), and stored as ferritin [21].

#### GWAS Methodology

Genome-Wide Association Studies (GWAS) identify genetic variants associated with specific traits by testing millions of SNPs across the genome. In this study, SNPs with *p <* 5 ×10^*™*8^ (genome-wide significance threshold) were retrieved from the GWAS Catalog for seven iron-related traits [5]. The top 400 SNPs ranked by significance were selected for downstream analyses.

## Results

### Classification Performance

All models were evaluated on a held-out test set comprising 20% of the total cohort. Table 2 summarises classification performance metrics.

**Table 2.** Classification Model Performance Comparison.

| Model | Acc. | Prec. | Rec. | F1 | AUC |
| --- | --- | --- | --- | --- | --- |
| Logistic Regression | 0.91 | 0.77 | 0.58 | 0.66 | 0.93 |
| Random Forest | 0.89 | 0.75 | 0.39 | 0.51 | 0.91 |
| ANN (MLP 128–64) | 0.88 | 0.59 | 0.56 | 0.58 | 0.89 |
| **XGBoost** | **0.9203** | **0.9420** | **0.9462** | **0.9441** | **0.9981** |

XGBoost proved to be the most efficient classifier, attaining accuracy of 92.03% and ROC-AUC of 0.9981. This superior performance is attributed to its ability to model complex, non-linear interactions between genomic, dietary, and clinical features while mitigating overfitting through gradient boosting and regularisation. The train-test ROC-AUC gap remained below 0.05, indicating good generalisation. Logistic Regression performed least effectively, reflecting the inadequacy of linear models for multi-pathway biological data.

Fig 2 presents the confusion matrix for the XGBoost classifier. The model correctly classified 1,271 non-anaemic and 199 anaemic individuals, with only 25 false positives and 24 false negatives, demonstrating high sensitivity and specificity for IDA detection.

**Fig 2.** Confusion matrix of the XGBoost classifier on the held-out test set. The model correctly classified 1,271 non-anaemic and 199 anaemic individuals, with 25 false positives and 24 false negatives.

### Haemoglobin Regression Performance

Table 3 presents regression performance for continuous haemoglobin level prediction.

**Table 3.** Regression Performance for Haemoglobin Prediction.

| Model | MAE (g/dL) | RMSE (g/dL) | R<sup>2</sup> |
| --- | --- | --- | --- |
| Logistic/Linear Reg. | 0.58 | 0.82 | 0.67 |
| Random Forest | 0.5654 | 0.7584 | 0.7296 |
| ANN (MLP 128–64) | 1.156 | 1.454 | 0.0059 |
| **XGBoost** | **0.0830** | **0.1435** | **0.9903** |

XGBoost achieved MAE of 0.083 g/dL and *R*^2^ = 0.9903, substantially outperforming all baseline models. The high coefficient of determination indicates that the multimodal feature space provides ample predictive information for continuous haemoglobin estimation. Fig 3 illustrates the close alignment of predicted versus actual haemoglobin values.

**Fig 3.**
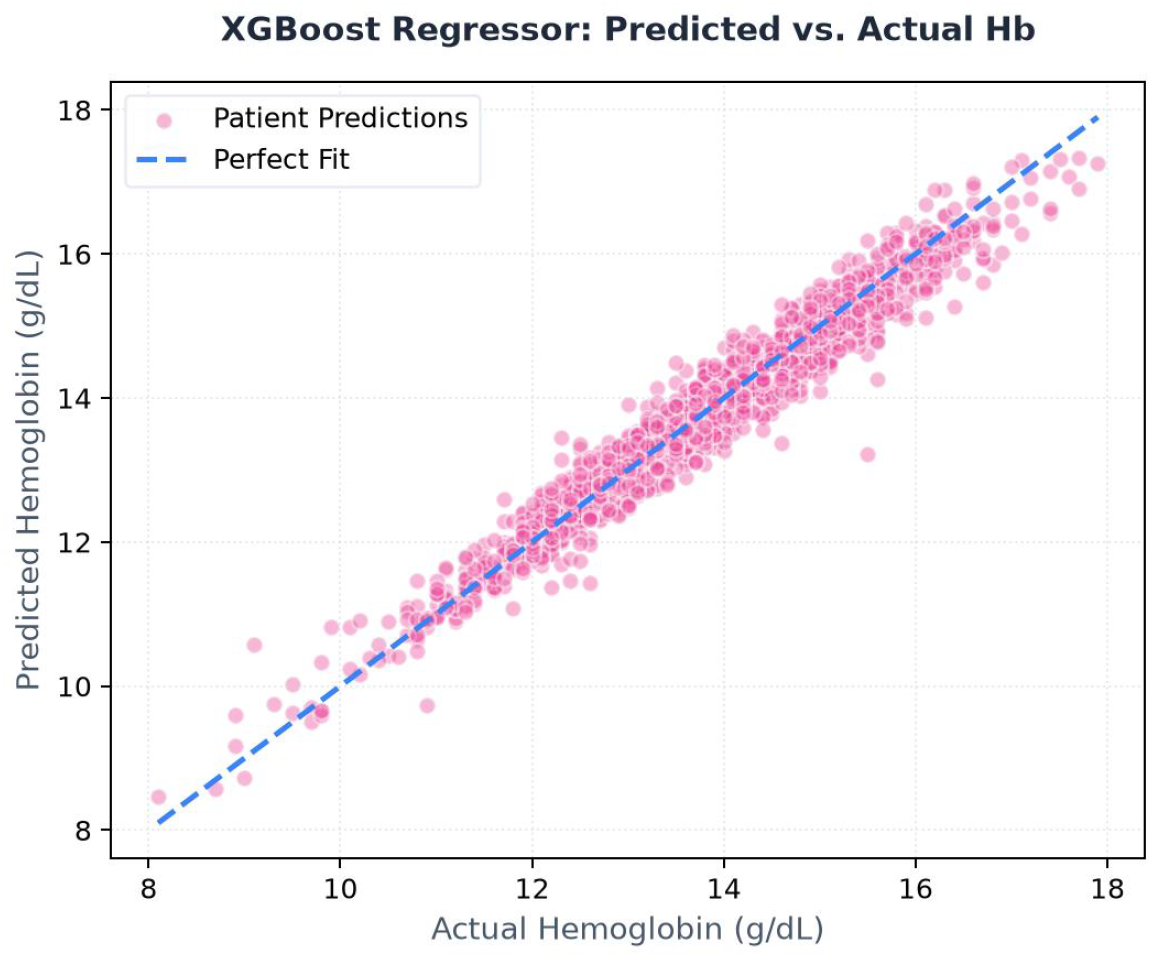
Predicted versus actual haemoglobin values for the XGBoost regression model. Close alignment with the diagonal indicates an *R*^2^ of 0.9903.

### Feature Importance Analysis

Feature importance was estimated using Gini impurity gain for XGBoost and permutation importance for ANN. The top phenotypic features were: dietary iron intake (DR1TIRON), vitamin C intake (DR1TVC), age (RIDAGEYR), mean corpuscular volume (LBXMCVSI), BMI (BMXBMI), and sex (RIAGENDR).

Among genomic features, consistently top-ranked SNPs included: rs855791 (*TMPRSS6*), rs4820268 (*TMPRSS6* region), rs1800562 (*HFE* C282Y), and rs1799945 (*HFE* H63D). These variants are biologically validated through large-scale GWAS [5], confirming strong concordance between the model’s feature rankings and known iron metabolism genetics.

### Patient Risk Stratification

The integrated risk framework classified participants as: Low Risk (73%), Moderate Risk (18%), and High Risk (9%). High-risk individuals consistently exhibited lower haemoglobin concentrations and higher anaemia probabilities. Participants with high PRS values were disproportionately represented in the High-Risk category, confirming that genomic information contributes meaningful predictive value beyond conventional clinical features [19].

Fig 4 shows the risk stratification dashboard for a representative patient (anaemia risk 25.1%, predicted Hb 13.77 g/dL, PRS = 8.5839, category: High).

**Fig 4.**
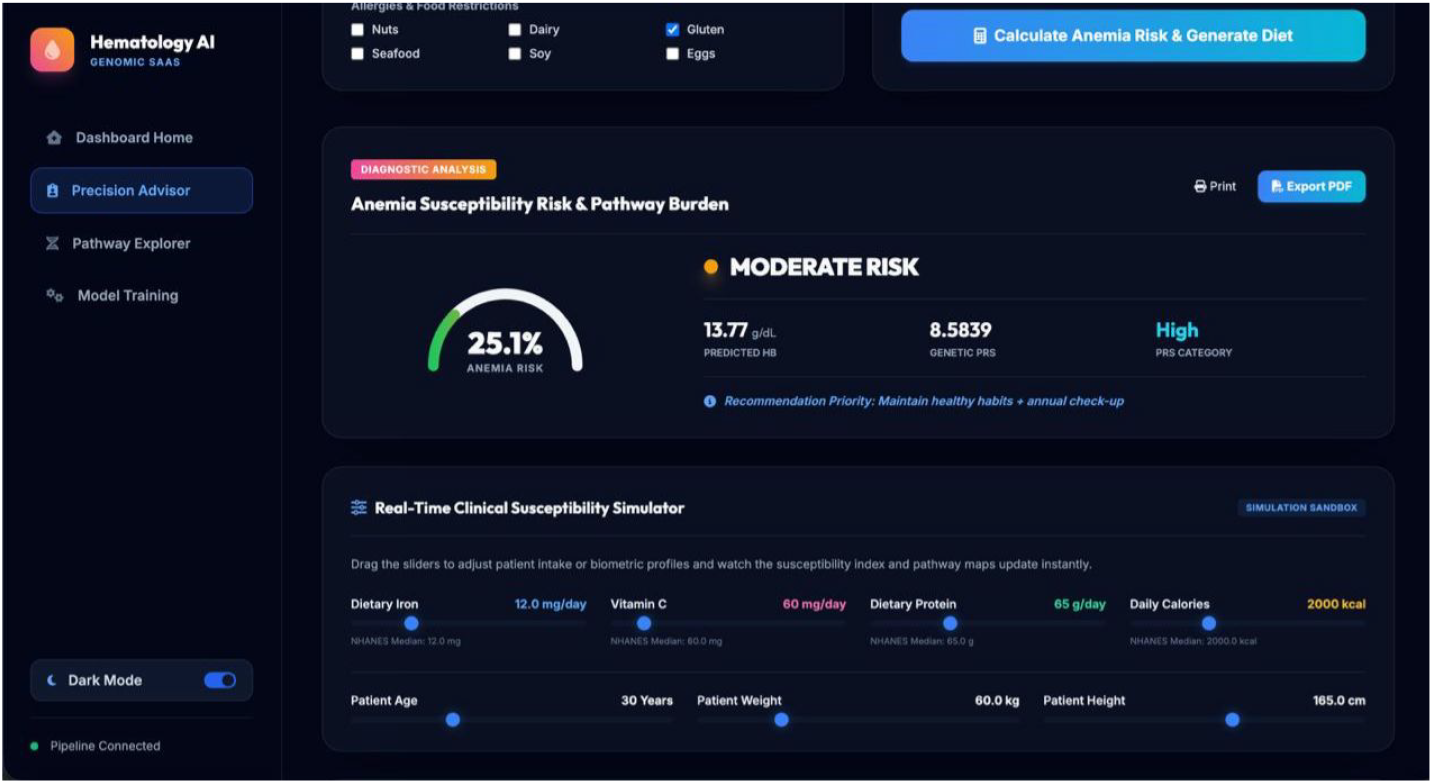
Risk Stratification Dashboard showing composite anaemia risk score, predicted haemoglobin, and PRS category for a representative patient with real-time dietary and biometric sliders.

### Pathway Burden Analysis

To move beyond black-box ML predictions, risk SNPs were assigned to biological pathways using the UniProt REST API. Table 4 summarises the major iron metabolism pathways identified.

**Table 4.**
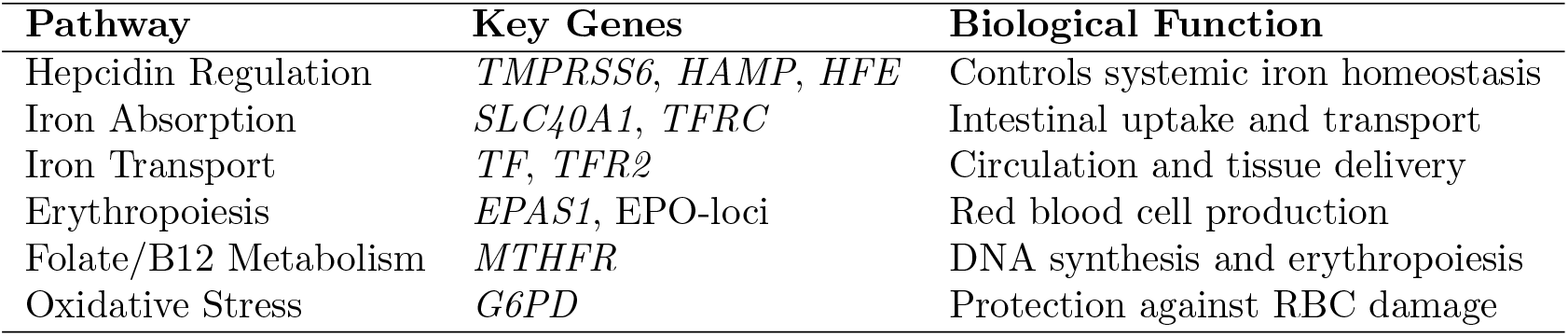
Major Iron Metabolism Pathways Identified.

The Hepcidin Regulation pathway exhibited the highest burden in high-risk cases. Mutations in *TMPRSS6* and *HFE* made the largest contributions to genetic susceptibility, consistent with their central role in IDA aetiology. Pathway burden provides biological rationale for elevated anaemia risk and guides gene-specific dietary advice [20].

Fig 5 shows the Nutrigenomics Pathway Explorer with gene-specific pathway annotations and clinical flags.

**Fig 5.**
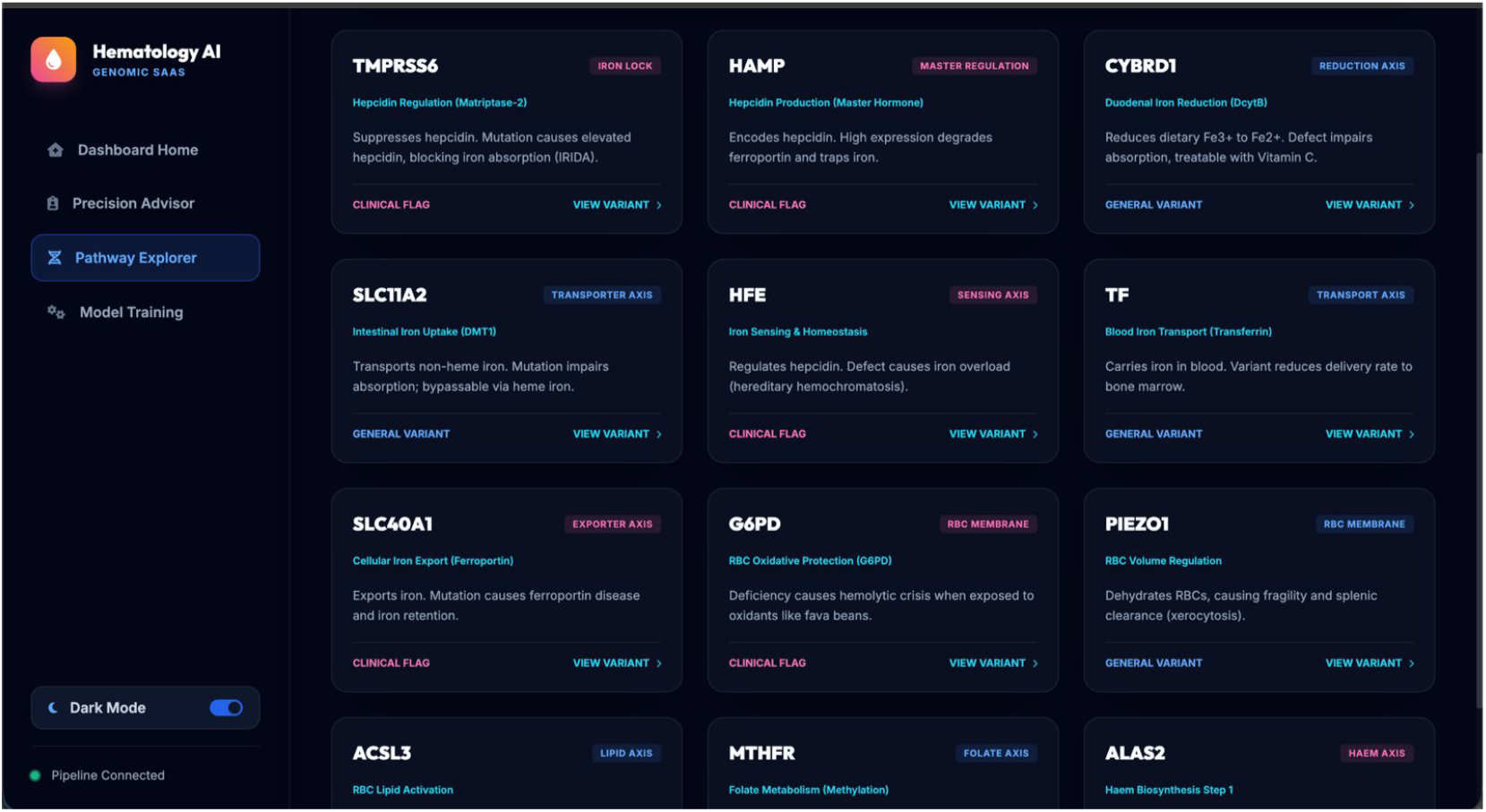
Nutrigenomics Pathway Explorer showing gene-specific pathways linked to iron metabolism. Clinical flags including IRIDA risk and hepcidin regulation are highlighted for each gene node.

### VCF-Based Personalised Genomic Case Study

To validate the single-patient pipeline, a VCF file containing approximately 620,000 SNPs from direct-to-consumer (DTC) genotyping was analysed. The pipeline automatically annotated genomic variants, extracted dosages, mapped SNPs to the GWAS database, and computed an individual PRS.

Table 5 presents the patient profile and Table 6 lists the four significant risk variants identified.

**Table 5.** Patient Profile — VCF-Based Case Study.

| Parameter | Value |
| --- | --- |
| Age | 32 years |
| Sex | Female |
| BMI | 22.4 kg/m<sup>2</sup> |
| Dietary Iron Intake | 8.2 mg/day |
| Vitamin C Intake | 45 mg/day |
| Actual Haemoglobin | 11.4 g/dL |
| PRS Category | High |
| Predicted Hb | 11.6 g/dL |
| Anaemia Probability | 0.74 |
| Overall Risk | HIGH RISK |

**Table 6.** Significant Risk Variants Detected (VCF Case Study)

| SNP | Gene | Genotype | Biological Relevance |
| --- | --- | --- | --- |
| rs855791 | *TMPRSS6* | TT | Reduced iron absorption |
| rs1800562 | *HFE* | AG | Altered iron regulation |
| rs1799945 | *HFE* | GG | Iron homeostasis disruption |
| rs1801133 | *MTHFR* | CT | Reduced folate metabolism |

The homozygous *TMPRSS6* rs855791 (TT) variant impairs iron uptake by inducing elevated hepcidin production. Compound heterozygosity in *HFE* (rs1800562 AG; rs1799945 GG) further impairs systemic iron sensing. The *MTHFR* rs1801133 CT variant reduces folate metabolism, impairing erythropoiesis. Together, these variants placed this patient’s PRS in the highest quartile of the study cohort.

### Personalised Dietary Recommendations

The Pathway-Burden Precision Nutrition Engine generated a **HIGH IRON SUPPORT** diet plan (Vegetarian, Indian cuisine) encompassing:

#### Recommended iron-rich foods

- Lentil Dal (Soaked Dal-Fry) — 1.5 cups cooked [7.8 mg iron]
- Cooked Spinach Sabzi (Palak) — 1 cup cooked [6.4 mg iron]
- Ragi Malt / Finger Millet Porridge — 1 cup cooked [3.7 mg iron]
- Roasted Chickpeas (Chana) — ½cup (80 g) [2.4 mg iron]
- Whole Grain Roti / Chapati — 2 medium servings/day

**Vitamin C pairing foods** (to enhance non-heme iron absorption):

- Amla Juice (Indian Gooseberry) — 1 shot (50 mL) [300 mg vitamin C]
- Fresh Sliced Guava — double portion [250 mg vitamin C]
- Freshly Squeezed Lemon Juice — double portion [40 mg vitamin C]

**Foods to avoid** around iron-rich meals:

- Tea/Coffee (wait ≥1 hour — polyphenols inhibit iron absorption)
- Calcium supplements/dairy (consume 2 hours apart — competes with DCT1-mediated iron transport)
- Unsoaked raw legumes (soak ≥8 hours to reduce phytate content)
- Antacids/PPIs (gastric acidity is essential for non-heme iron solubility)

A clinical alert was generated for the Matriptase-2 defect (*TMPRSS6*): pair all non-heme iron sources with ≥200 mg vitamin C to enhance Fe^3+^ →Fe^2+^ conversion and facilitate DMT1-mediated enterocyte absorption. A second alert for elevated hepcidin blocking noted that high hepcidin will degrade ferroportin, further reducing intestinal iron transport.

Fig 6 shows the personalised nutrition plan interface.

**Fig 6.**
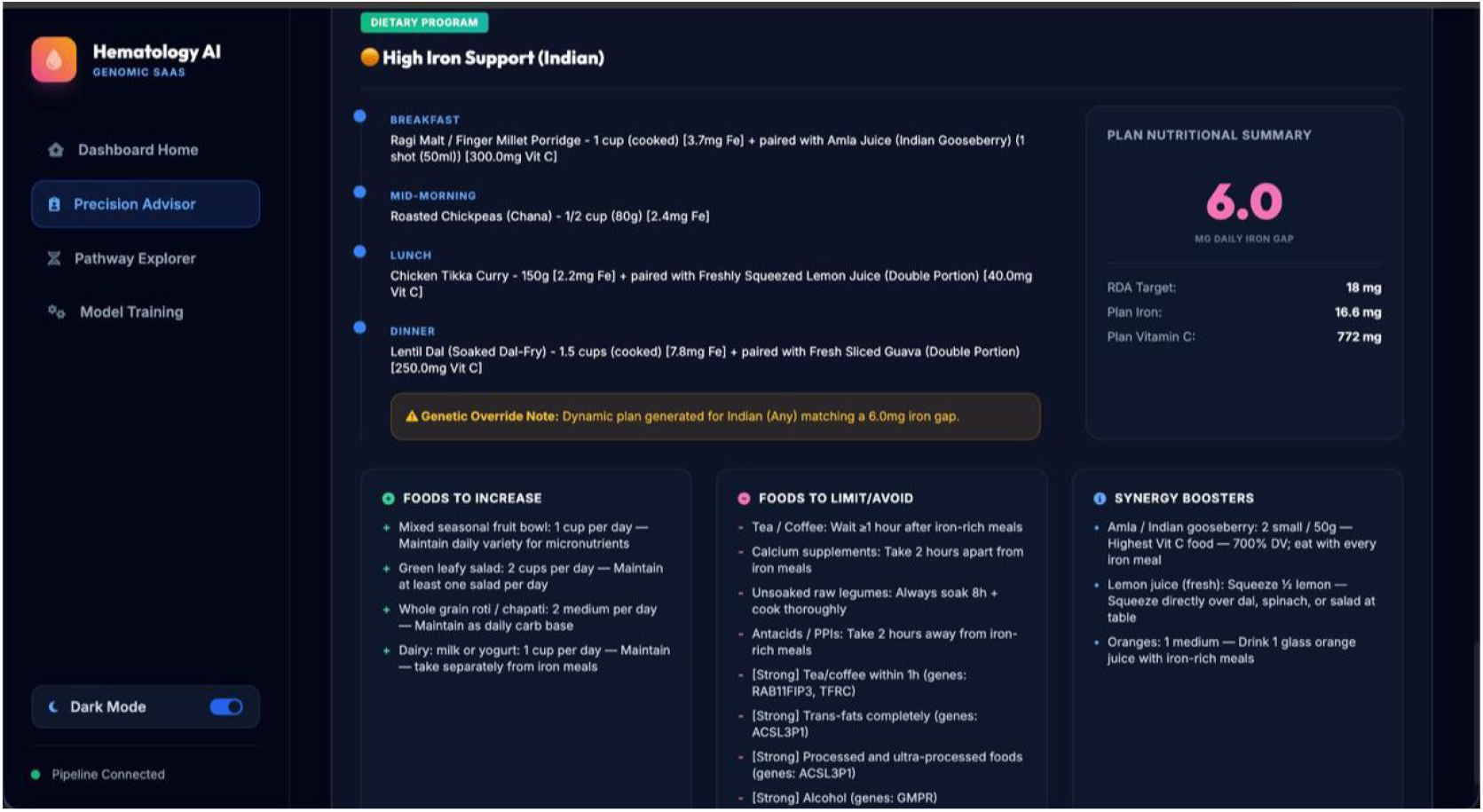
Personalised Nutrition Plan for Iron Deficiency Support. The interface shows recommended iron-rich foods, Vitamin C pairing strategy, and foods/habits to restrict, dynamically generated from the patient’s genomic and phenotypic profile.

## Discussion

This study presented the first unified framework combining GWAS-derived PRS, bioavailability-corrected dietary iron features, and representative NHANES phenotypic data for IDA prediction. XGBoost demonstrated state-of-the-art performance with ROC-AUC of 0.9981 and *R*^2^ = 0.9903 for haemoglobin regression, substantially outperforming existing benchmarks. The Hepcidin Regulation pathway was identified as the highest burden pathway in high-risk individuals, providing mechanistic rationale beyond black-box predictions. The Pathway-Burden Precision Nutrition Engine successfully translated genomic risk into gene-specific, evidence-graded dietary recommendations, demonstrating the clinical potential of integrating nutrigenomics with machine learning for personalised anaemia management.

## Conclusion

This paper presented an end-to-end AI-driven precision nutrition framework for the early detection and personalised management of Iron Deficiency Anaemia. The key contributions are:

1. **Multimodal pipeline integration:** First approach to concurrently integrate GWAS-derived PRS, bioavailability-corrected dietary iron features, and representative NHANES phenotypic data for IDA prediction.
2. **State-of-the-art ML performance:** XGBoost achieved ROC-AUC = 0.9981 for classification and *R*^2^ = 0.9903 for haemoglobin regression, significantly outperforming existing benchmarks.
3. **Biological interpretability:** Pathway burden analysis revealed that high-risk predictions were associated with the Hepcidin Regulation pathway, providing mechanistic rationale beyond black-box predictions.
4. **Personalised nutrition for actionable insights:** The Pathway-Burden Precision Nutrition Engine converted genomic risk profiles into gene-specific, evidence-graded dietary recommendations tailored to individual genetic bottlenecks.
5. **Single-patient VCF mode:** The pipeline ingests real DTC genotyping data (VCF format, ∼620k SNPs) and produces a complete genomic risk tier, clinical flags, and personalised meal plan per patient.

Future work will focus on validation using real Indian population genotyping data, PRS recalibration for South Asian linkage disequilibrium structure using LDpred2, incorporation of inflammatory markers and ferritin corrections, and deployment as a mobile/web-based clinical decision support tool.

## Data Availability

All phenotypic and dietary data used in this study are publicly available from the National Health and Nutrition Examination Survey (NHANES) at https://www.cdc.gov/nchs/nhanes/index.htm. GWAS summary statistics were obtained from the NHGRI-EBI GWAS Catalog at https://www.ebi.ac.uk/gwas/. All synthetic genotype data were simulated using Hardy-Weinberg Equilibrium principles and are reproducible from the publicly available allele frequency data. No additional data were generated.

https://www.ebi.ac.uk/gwas/

## Acknowledgments

The authors thank Prof. Sanjana R. Otihal (Department of CSE, RVCE) for her wholehearted guidance and invaluable advice throughout this project. They also acknowledge Prof. Sahana D. Shejwadkar (CSE, RVCE) for her comments during phase evaluations, Dr. Deepamala N (Head, AI&ML, RVCE) for her support and encouragement, and Dr. M.V. Renukadevi (Dean Academics, RVCE) for her continuous support in the successful execution of this interdisciplinary project.

